# Japanese citizens’ behavioral changes and preparedness against COVID-19: How effective is Japan’s approach of self-restraint ?

**DOI:** 10.1101/2020.03.31.20048876

**Authors:** Kaori Muto, Isamu Yamamoto, Miwako Nagasu, Mikihito Tanaka, Koji Wada

## Abstract

The Japanese government instituted countermeasures against COVID-19, a pneumonia caused by the new coronavirus, in January 2020. Seeking “people’s behavioral changes,” in which the government called on the public to take precautionary measures or exercise self-restraint, was one of the important strategies. The purpose of this study is to investigate how and from when Japanese citizens have changed their precautionary behavior under these circumstances, where the government has only requested their cooperation. This study uses micro data from a cross-sectional survey conducted on an online platform of an online research company, based on quota sampling that is representative of the Japanese population. By the end of March 2020, we had recruited a total of 11,342 respondents, aged from 20 to 64 years. About 85% reported practising the social distancing recommended by the government. More females than males and more older than younger participants are supportive of practicing social distancing. Frequent handwashing is conducted by 86 percent of all, 92 percent of female and 87.9 percent of over-40 participants. The most important event influencing these precautionary actions was the infection aboard the Diamond Princess cruise ship, which occurred in early February 2020 (23%). Information from the central and local governments, received by 60% of the participants, was deemed trustworthy by 50%. However, the results also showed that about 20% of the participants were reluctant to implement proper prevention measures. The statistical analysis indicated that the typical characteristics of those people were male, younger (under 30 years old), unmarried, from lower-income households, with a drinking or smoking habit and a higher extraversion score. To prevent the spread of infection in Japan, it is imperative to address these individuals and encourage their behavioral changes using various means to reach and influence them.

## Introduction

### The new coronavirus in Japan

Pneumonia of unknown cause was detected in China and first officially reported on 31 December 2019. The World Health Organization (WHO) announced a name for the new coronavirus disease, COVID-19 (coronavirus disease 2019), on 11 February 2020.^[1]^ Since then, COVID-19 has been spreading throughout the world, and a rapid increase in deaths has been reported in many countries. As of 28 March, a total of 571,678 cases and 26,494 deaths have been confirmed.^[2]^ One study has estimated that there will be a total of 81,114 deaths from COVID-19 over the next four months in the US alone.^[3]^ The number of COVID-19 cases and deaths in Japan is gradually increasing, with 1,499 cases (including 60 critical cases) and 49 deaths reported as of 28 March.^[4]^ Several small clusters of infected groups have been increasing in urban areas, including those in hospitals and nursing homes, in addition to cases with unlinked infections. Nevertheless, the total number of deaths and severely ill patients has been comparatively small, especially relative to the country’s population size, and the trend of the increase is not sharp. The reasons for this mild trend have been questioned outside Japan.^[5]^

Over the past few decades, Japan has not experienced any serious damage from new infectious diseases, such as SARS (severe acute respiratory syndrome), MERS (Middle East respiratory syndrome) or the Ebola virus. Ironically, this history of escapes might delay the establishment of the emergency operation headquarters in Japan. The implementation of polymerase chain reaction (PCR) tests, which must be the frontline response to the novel coronavirus outbreak, has faced time-consuming obstacles. In Japan, a recent revision of the Act on Special Measures for Pandemic Influenza and New Infectious Diseases Preparedness and Response allows the Prime Minister to declare a state of emergency for the outbreak, but under the current legislation, no central or local government can enforce lockdowns such as those undertaken in other countries. If the Act on the Prevention of Infectious Diseases and Medical Care for Patients with Infectious Diseases is revised for COVID-19, local governors can restrict or block the traffic in contaminated places for a specified period of not more than 72 hours for the purpose of preventing the spread of the coronavirus.

Under such limitations, the current goal of the Japanese government is to avoid an explosive increase in patients that would exceed the limit of intensive or critical care units in hospitals in urban areas. To meet this goal, the government policy consists of three strategies: early detection of clusters and rapid response, enhancement of the early diagnosis of patients and intensive care for severely affected patients, and strengthening of the universal healthcare system and people’s behavioral change.^[6]^

### Three strategies against COVID-19

With regards to the first and second strategies, the Ministry of Health, Labor and Welfare (MHLW) strongly promotes contact tracing, social distancing and pneumonia surveillance under the direction of the Patient Cluster Countermeasure Group in the MHLW Headquarters for Novel Coronavirus Disease Control. Regional public health centers conduct contact tracing, asking infected persons and their close contacts to maintain social distancing for 14 days and allocating available hospital beds or hospital wards in designated local communities. In clinical settings, the large number of computed tomography (CT) scanners in Japan (111.49 per million population^[7]^) supports physicians with investigating suspicious pneumonia cases in the absence of conducting massive PCR tests in the population. This policy approach might lead to a relatively slower increase in the number of cases and deaths.

Regarding the third strategy, people’s behavioral change, by the middle of February 2020, the MHLW encouraged the Japanese public to practise frequent handwashing and “coughing etiquette” (using a handkerchief or sleeve instead of hands to catch a cough or sneeze). Furthermore, the MHLW has prioritized access to healthcare for elderly people, people suffering from fatigue or shortness of breath and people with underlying health conditions. The MHLW has also asked the public not to visit clinics for at least four days if they have experienced cold symptoms or a fever of 37.5°C or over.^[8]^ This restriction might be a shock to Japanese citizens, who are typically allowed to freely access clinics and hospitals.

In analyses of contact tracing, it was found that one infected person tended to infect more than one other person at locations with certain characteristics. On 24 February, the Expert Meeting on the Control of Novel Coronavirus Infection asked the public to refrain from attending places involving close face-to-face contact (within an arm’s length of each other) in conversations and similar interactions for more than a given length of time in crowds. Since then but prior to other similar slogans that have appeared in the world, the government has been campaigning for avoidance of these situations with the slogan “Avoid overlapping three Ms” *mippei-kukan* [poorly ventilated closed space], *misshu-basho* [large gathering] and *missetsu-bamen* [conversations or shouting in close proximity], in addition to regular ventilation and wiping of shared surfaces (door handles, knobs, and bed fences) and goods with diluted household chlorine bleach. “Avoid overlapping three Ms” has been the core and unique message against COVID-19 in Japan.

### Previous studies and our research questions

This study examines three research questions: 1) How do Japanese citizens implement the government’s three Ms precautionary measures? 2) How effective are these requests from the government? and 3) Who has changed their daily precautionary behavior and who has not?

Several previous studies have investigated changes in people’s precautionary behavior against the coronavirus. For example, an online survey conducted on 29 January of 3,083 mainland Chinese respondents revealed that adults living in urban areas had a better awareness of the issue than those in rural areas (72.7% vs. 66.1%, p<0.001).^[9]^ Another online survey conducted between 23 February and 2 March in the US (N=2,986) and the UK (N=2,988) adult residents showed that people have a good understanding of the main mode of disease transmission and common symptoms, although they also have several important misconceptions and discriminatory attitudes towards people with East Asian ethnicity due to COVID-19’s origin in China.^[10]^ The latest study in Italy clarified the three types of attitude to COVID-19 among Italian citizens: people who trust authority and choose isolation, fatalists who are keen on social media and uniformed youth. ^[11]^ The Gallup International Association also recently conducted a snap poll in 28 countries (including 1,115 Japanese participants), asking about precautionary procedures, and their findings indicated that 71 percent of Japanese participants had adopted more frequent handwashing.^[12]^ It is still unclear, however, what the trigger is for behavioral change around COVID-19 and who is more actively implementing the prevention measures. In this survey, the response period and sample attribution are also unclear. Furthermore, this survey is not necessarily informative for policymaking, as it does not reveal who is *not* implementing prevention measures.

Using a large sample of cross-sectional survey data, this study investigates how and at what point Japanese citizens changed their precautionary behavior in this situation, where the government has only requested, rather than mandated, their cooperation.

## Materials and Methods

### Survey design and participants

This study uses micro data from a cross-sectional survey conducted via an online platform of an online research company, MACROMILL INC, Japan. From a pool of approximately 1.2 million registered individuals residing in Japan, we recruited a total of 11,342 males and females aged from 20 to 64 years. In the recruitment process for this study, quota sampling was conducted so that the sample distributions among gender (male or female), age group (20s, 30s, 40s, 50s or 60s), and employment status (regular employee, non-regular employee, self-employed or not working) became equal to those of the representative Japanese population, based on the statistics of the Labor Force Survey (Ministry of Internal Affairs and Communications). The survey was conducted between 26 and 28 March 2020.

### Questionnaire and analysis

In addition to providing individual characteristics, the participants were asked to answer 11 items rating their prevention measures against novel coronavirus infections, such as social distancing and coughing etiquette on a scale of 1 to 5. Thus, after summarizing demographic characteristics based on the total, male and female, and under-40 and over-40 categories, we aggregate and compare a proportion of the participants who have been taking those prevention measures.

The participants were also asked what kind of events caused them to change their behaviors and rated the frequency and reliability of 10 information sources about the coronavirus on a scale of 1 to 5. Thus, we calculate and compare the frequency and reliability depending on the information sources.

Next, to detect factors associated with behavioral change, the participants were also asked about their drinking and smoking habits. Personality traits were measured by the Five Factor Personality Questionnaire: Ten-Item Personality Inventory (TIPI). We estimate a logit model, where the dependent variable is a dummy indicating 1 if the participant chose “not at all” or “not true” to the question “Do you avoid the three overlapping Ms?” and where independent variables are individual characteristics.

### Data analysis

We analyzed the data using STATA/MP version 16.0 for Mac (StataCorp, College Station, TX, United States).

### Ethical issues

Our survey falls outside the scope of the Japanese government’s Ethical Guidelines for Medical and Health Research Involving Human Subjects, and there are no national guidelines in Japan for social and behavioral research. Therefore, our study was carried out in accordance with the Ethical Principles for Sociological Research of the Japan Sociological Society, which do not require ethical reviews.

All survey participants gave consent to participate in the anonymous online survey by MACROMILL INC. The authors did not obtain any personal information about the participants. After being informed about the purposes of the study and their right to quit the survey, participants agreed to participate. They were provided with the option of “I don’t want to respond” for all questions. Completion of the entire questionnaire was considered as participant consent.

## Results

### Demographic characteristics

The characteristics of the sample, both as a whole and separated by gender (male or female) or age (under or over 40 years old), are summarized in Table 1. The total sample size is 11,342, with almost equal gender distribution. Gender and age distribution are proportional to that of the Japanese population. University or college graduates constituted about 50–60 percent of respondents. About half of the total sample is composed of regular employees (usually, indefinite and full-time employees). About a quarter of respondents had a household income of 4–5 million yen.

**Table 1.** **Sample characteristics**

|  | n, (%) |  |  |  |  |  |  |  |  |
| --- | --- | --- | --- | --- | --- | --- | --- | --- | --- |
|  | All |  | Male |  | Female |  | Under 40 y |  | Over 40 y |
| Total respondents | 11,342 |  | 5,734 |  | 5,608 |  | 4,300 |  | 7,842 |
| Female | 5,608 (49.44) |  |  |  |  |  | 2,110 (49.07) |  | 3,498 (49.67) |
| Age |  |  |  |  |  |  |  |  |  |
| 20-29 years old | 1,964 (17.32) |  | 994 (17.34) |  | 970 (17.30) |  | 1,964 (45.67) |  | 0 (0.00) |
| 30-39 years old | 2,336 (20.60) |  | 1,196 (20.86) |  | 1,140 (20.33) |  | 2,336 (54.33) |  | 0 (0.00) |
| 40-49 years old | 3,098 (27.31) |  | 1,568 (27.35) |  | 1,530 (27.28) |  | 0 (0.00) |  | 3,098 (43.99) |
| 50-59 years old | 2,754 (24.28) |  | 1,373 (23.94) |  | 1,381 (24.63) |  | 0 (0.00) |  | 2,754 (39.11) |
| 60-64 years old | 1,190 (10.49) |  | 603 (10.52) |  | 587 (10.47) |  | 0 (0.00) |  | 1,190 (16.90) |
| Married | 6,620 (58.37) |  | 3,241 (56.52) |  | 3,379 (60.25) |  | 1,859 (43.23) |  | 4,761 (67.61) |
| Parents with children under junior high school | 2,972 (26.20) |  | 1,493 (26.04) |  | 1,479 (26.37) |  | 1,464 (34.05) |  | 1,508 (21.41) |
| University or college graduate | 6,278 (55.35) |  | 3,415 (59.56) |  | 2,863 (51.05) |  | 2,478 (57.63) |  | 3,800 (53.96) |
| Work status |  |  |  |  |  |  |  |  |  |
| Regular employee | 5,817 (51.29) |  | 3,986 (69.52) |  | 1,831 (32.65) |  | 2,460 (57.21) |  | 3,357 (47.67) |
| Nonregular employee | 2,865 (25.26) |  | 733 (12.78) |  | 2,132 (38.02) |  | 1,010 (23.49) |  | 1,855 (26.34) |
| Self-employed and others | 660 (5.82) |  | 422 (7.36) |  | 238 (4.24) |  | 138 (3.21) |  | 522 (7.41) |
| Not working | 2,000 (17.63) |  | 593 (10.34) |  | 1,407 (25.09) |  | 692 (16.09) |  | 1,308 (18.57) |
| Household annual income |  |  |  |  |  |  |  |  |  |
| Less than 2,000K JPY | 646 (7.56) |  | 312 (6.67) |  | 334 (8.63) |  | 250 (8.28) |  | 396 (7.16) |
| 2,000-3,999K JPY | 1,939 (22.68) |  | 925 (19.77) |  | 1,014 (26.21) |  | 773 (25.59) |  | 1,166 (21.10) |
| 4,000-5,999K JPY | 2,247 (26.29) |  | 1,224 (26.16) |  | 1,023 (26.44) |  | 869 (28.77) |  | 1,378 (24.93) |
| 6,000-6,999K JPY | 1,606 (18.79) |  | 929 (19.85) |  | 677 (17.50) |  | 541 (17.91) |  | 1,065 (19.27) |
| 8,000-8,999K JPY | 1,052 (12.31) |  | 619 (13.23) |  | 433 (11.19) |  | 322 (10.66) |  | 730 (13.21) |
| 10,000-11,999K JPY | 516 (6.04) |  | 318 (6.80) |  | 198 (5.12) |  | 139 (4.60) |  | 377 (6.82) |
| 12,000-14,999K JPY | 306 (3.58) |  | 192 (4.10) |  | 114 (2.95) |  | 58 (1.92) |  | 248 (4.49) |
| 15,000-19,999K JPY | 151 (1.77) |  | 100 (2.14) |  | 51 (1.32) |  | 42 (1.39) |  | 109 (1.97) |
| More than 20,000K JPY | 85 (0.99) |  | 60 (1.28) |  | 25 (0.65) |  | 27 (0.89) |  | 58 (1.05) |

### To what extent have prevention measures been taken?

In the survey, the participants were first asked to answer to the question “Have you ever conducted any measures to prevent novel coronavirus infections or outbreaks?” About 76 percent of participants evaluated themselves as having taken some action.

To observe the details of those actions, Table 2 shows a variety of prevention measures taken, aggregating a proportion of the participants who answer “very true” and “true” for each prevention measure.

**Table 2.** **“Have you ever conducted anything to prevent novel coronavirus infections or outbreaks?”**

|  | % of very true and true, (C.I.) |  |  |  |  |
| --- | --- | --- | --- | --- | --- |
|  | All | Male | Female | Under 40 y | Over 40 y |
| 1. Avoid a poorly-ventilated closed space | 80.6<br>(79.8 - 81.3) | 75.9<br>(74.8 - 77.0) | 85.3<br>(84.4 - 86.3) | 77.6<br>(76.3 - 78.8) | 82.4<br>(81.5 - 83.3) |
| 2. Avoid large gatherings | 80.5<br>(79.8 - 81.2) | 77.5<br>(76.4 - 78.6) | 83.6<br>(82.6 - 84.6) | 76.3<br>(75.1 - 77.6) | 83.1<br>(82.2 - 83.9) |
| 3. Avoid conversations or shouting in close proximity | 57.0<br>(56.1 - 57.9) | 55.6<br>(54.3 - 56.8) | 58.5<br>(57.2 - 59.8) | 52.3<br>(50.9 - 53.8) | 59.9<br>(58.7 - 61.0) |
| 4. Avoid places where items 1-3 above overlap | 80.6<br>(79.8 - 81.3) | 76.9<br>(75.8 - 77.9) | 84.4<br>(83.4 - 85.3) | 76.7<br>(75.4 - 77.9) | 82.9<br>(82.1 - 83.8) |
| 5. Do not go to mass gatherings | 86.8<br>(86.2 - 87.5) | 82.7<br>(81.8 - 83.7) | 91.0<br>(90.3 - 91.8) | 82.7<br>(81.6 - 83.8) | 89.4<br>(88.6 - 90.1) |
| 6. Undertake frequent handwashing | 86.3<br>(85.7 - 87.0) | 81.9<br>(80.9 - 82.9) | 90.9<br>(90.2 - 91.7) | 83.8<br>(82.7 - 84.9) | 87.9<br>(87.1 - 88.6) |
| 7. Undertake cough etiquette (use handkerchiefs or sleeves instead of hands) | 77.0<br>(76.2 - 77.8) | 72.0<br>(70.9 - 73.2) | 82.1<br>(81.1 - 83.1) | 73.9<br>(72.6 - 75.2) | 78.9<br>(77.9 - 79.8) |
| 8. Always wear a surgical-style mask when going out | 70.1<br>(69.2 - 70.9) | 62.7<br>(61.4 - 63.9) | 77.6<br>(76.5 - 78.7) | 70.0<br>(68.6 - 71.3) | 70.1<br>(69.1 - 71.2) |
| 9. Avoid going out when you have a cold | 76.7<br>(76.0 - 77.5) | 70.9<br>(69.7 - 72.1) | 82.7<br>(81.7 - 83.7) | 72.8<br>(71.4 - 74.1) | 79.2<br>(78.2 - 80.1) |
| 10. Get sufficient rest and sleep | 73.1<br>(72.3 - 73.9) | 68.3<br>(67.1 - 69.5) | 77.9<br>(76.9 - 79.0) | 71.6<br>(70.2 - 72.9) | 74.0<br>(73.0 - 75.0) |
| 11. Eat a nutritious diet | 69.5<br>(68.6 - 70.3) | 64.2<br>(62.9 - 65.4) | 74.9<br>(73.7 - 76.0) | 69.1<br>(67.7 - 70.5) | 69.7<br>(68.6 - 70.8) |
| 12. Prepare consultation and transportation methods for when you feel ill | 41.5<br>(40.6 - 42.4) | 42.7<br>(41.4 - 44.0) | 40.3<br>(39.0 - 41.6) | 41.9<br>(40.4 - 43.3) | 41.3<br>(40.1 - 42.4) |

Looking at the first four prevention measures, which have been continuously requested by the Japanese government and the Expert Meeting on Control of Novel Coronavirus Infection, it was found that 80 percent have attempted to avoid the “overlapping three Ms.” Of the total, 57% have attempted avoid conversations or shouting in close proximity, which was a relatively low figure among the three Ms. Looking next at the fifth prevention measure, more than 85 percent of all participants reported practising social distancing by avoiding mass gatherings. Regarding gender and age differences, more females than males and more older than younger participants are supportive of social distancing, as shown by the differences in the confidence intervals.

Regarding hygiene practices, frequent handwashing is conducted by about 86 percent of all, about 91 percent of female and about 88 percent of over-40 participants. Coughing etiquette was implemented by 77 percent of the participants. Many also answered that they have avoided going out when ill with a cold.

As for the measures to strengthen individual immunity, around 70 percent of the participants reported getting sufficient rest and sleep or eating a nutritious diet. Again, focusing on gender and age differences, prevention measures are conducted more often by females and older people.

However, regardless of gender and age, about 40 percent of participants have prepared consultation and transportation methods for if they were to become ill.

### What has caused the behavioral changes?

To explore the triggers of the behavioral changes and preparedness observed above, the participants were asked “What was the most important event influencing these actions?”; the responses are summarized in Fig 1. The figure shows that about 23 percent of the participants cited the infection aboard the *Diamond Princess* cruise ship ^[13]^ that occurred around early February 2020, when there were still few domestic cases. *Diamond Princess* is a British-registered cruise ship on which an 80-year-old passenger from Hong Kong tested positive for COVID-19 on 1 February 2020. Because the ship was in Japanese waters, the ship was quarantined in February 2020 for nearly a month with about 3,700 passengers and crew on board. Other participants noted events from the end of February, including the alert from the Expert Meeting (5.6 percent), the statement of emergency by the governor of Hokkaido (northern island of Japan) (7.4 percent) and the request by the Prime Minister to not attend mass gatherings (7.8 percent). The next large trigger was the request by the Prime Minister for nationwide school closures in Japan on 28 February 2020 (about 14 percent). Finally, worldwide outbreak around early March (22 percent) also attracted participants’ attention.

**Fig 1.**
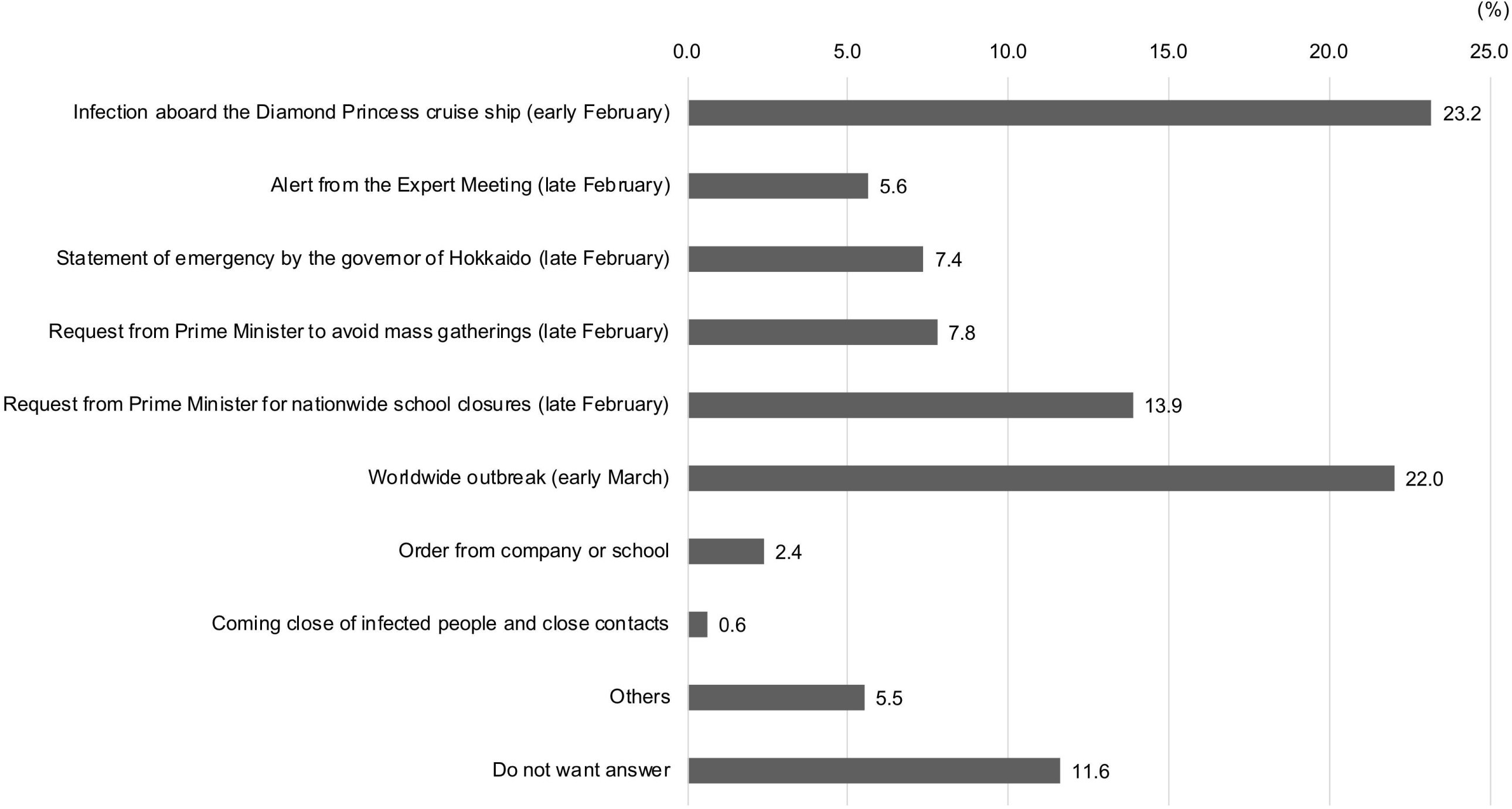
“What was the most important event influencing these actions?”

To explore what kind of information affected their behavioral change and preparedness, the survey asked participants to report the frequency at which they consult certain sources about the novel coronavirus infection and rate their reliability of the information source. The results are summarized in Table 3.

**Table 3.** **“From where do you get and trust information about novel coronavirus infection?”**

|  | %, (C.I.) |  |  |  |  |
| --- | --- | --- | --- | --- | --- |
|  | All | Male | Female | Under 40 y | Over 40 y |
| **1. TV news programs** |  |  |  |  |  |
| Get information: frequently or sometimes | 89.0<br>(88.5 - 89.6) | 85.4<br>(84.5 - 86.3) | 92.7<br>(92.1 - 93.4) | 85.2<br>(84.1 - 86.2) | 91.4<br>(90.8 - 92.1) |
| Trust information: very much or yes | 55.2<br>(54.3 - 56.1) | 51.0<br>(49.7 - 52.3) | 59.5<br>(58.3 - 60.8) | 50.5<br>(49.0 - 52.0) | 58.1<br>(56.9 - 59.2) |
| **2. TV talk and variety shows** |  |  |  |  |  |
| Get information: frequently or sometimes | 69.4<br>(68.6 - 70.3) | 62.4<br>(61.2 - 63.7) | 76.6<br>(75.5 - 77.7) | 67.5<br>(66.1 - 68.9) | 70.6<br>(69.5 - 71.7) |
| Trust information: very much or yes | 31.4<br>(30.6 - 32.3) | 27.2<br>(26.0 - 28.3) | 35.8<br>(34.5 - 37.0) | 32.5<br>(31.1 - 33.9) | 30.8<br>(29.7 - 31.9) |
| **3. Newspapers (national and local newspapers)** |  |  |  |  |  |
| Get information: frequently or sometimes | 42.0<br>(41.1 - 42.9) | 45.4<br>(44.1 - 46.7) | 38.6<br>(37.3 - 39.9) | 30.0<br>(28.6 - 31.4) | 49.4<br>(48.2 - 50.5) |
| Trust information: very much or yes | 47.5<br>(46.6 - 48.5) | 45.2<br>(43.9 - 46.4) | 50.0<br>(48.7 - 51.3) | 41.0<br>(39.6 - 42.5) | 51.5<br>(50.3 - 52.7) |
| **4. Tabloid paper** |  |  |  |  |  |
| Get information: frequently or sometimes | 7.9<br>(7.4 - 8.4) | 11.4<br>(10.5 - 12.2) | 4.3<br>(3.7 - 4.8) | 9.6<br>(8.7 - 10.5) | 6.8<br>(6.2 - 7.4) |
| Trust information: very much or yes | 12.7<br>(12.1 - 13.3) | 13.4<br>(12.6 - 14.3) | 11.9<br>(11.1 - 12.8) | 16.1<br>(15.0 - 17.2) | 10.6<br>(9.9 - 11.4) |
| **5. Internet news sites** |  |  |  |  |  |
| Get information: frequently or sometimes | 86.6<br>(86.0 - 87.3) | 84.7<br>(83.7 - 85.6) | 88.7<br>(87.8 - 89.5) | 84.4<br>(83.3 - 85.5) | 88.0<br>(87.3 - 88.8) |
| Trust information: very much or yes | 41.8<br>(40.9 - 42.7) | 41.3<br>(40.0 - 42.5) | 42.3<br>(41.0 - 43.6) | 42.1<br>(40.6 - 43.5) | 41.6<br>(40.5 - 42.8) |
| **6. SNS app news** |  |  |  |  |  |
| Get information: frequently or sometimes | 45.6<br>(44.7 - 46.5) | 42.8<br>(41.5 - 44.1) | 48.4<br>(47.1 - 49.8) | 57.5<br>(56.0 - 58.9) | 38.3<br>(37.2 - 39.5) |
| Trust information: very much or yes | 24.5<br>(23.7 - 25.3) | 23.4<br>(22.3 - 24.5) | 25.7<br>(24.5 - 26.8) | 29.9<br>(28.5 - 31.3) | 21.2<br>(20.3 - 22.2) |
| **7. Information sent by the Prime Minister** |  |  |  |  |  |
| Get information: frequently or sometimes | 66.3<br>(65.4 - 67.2) | 63.6<br>(62.3 - 64.8) | 69.1<br>(67.9 - 70.3) | 59.8<br>(58.3 - 61.3) | 70.3<br>(69.2 - 71.4) |
| Trust information: very much or yes | 47.5<br>(46.6 - 48.4) | 46.0<br>(44.8 - 47.3) | 48.9<br>(47.6 - 50.2) | 45.7<br>(44.2 - 47.2) | 48.6<br>(47.4 - 49.7) |
| **8. Information sent by the Ministry of Health, Labor and Welfare** |  |  |  |  |  |
| Get information: frequently or sometimes | 63.4<br>(62.5 - 64.3) | 60.8<br>(59.5 - 62.1) | 66.0<br>(64.8 - 67.2) | 58.3<br>(56.8 - 59.7) | 66.5<br>(65.4 - 67.6) |
| Trust information: very much or yes | 48.8<br>(47.9 - 49.7) | 46.9<br>(45.7 - 48.2) | 50.7<br>(49.4 - 52.0) | 47.6<br>(46.1 - 49.1) | 49.5<br>(48.4 - 50.7) |
| **9. Information provided by government Expert Meetings** |  |  |  |  |  |
| Get information: frequently or sometimes | 56.9<br>(55.9 - 57.8) | 55.2<br>(54.0 - 56.5) | 58.5<br>(57.2 - 59.8) | 50.3<br>(48.8 - 51.8) | 60.8<br>(59.7 - 62.0) |
| Trust information: very much or yes | 51.4<br>(50.5 - 52.3) | 49.7<br>(48.4 - 51.0) | 53.1<br>(51.8 - 54.4) | 48.8<br>(47.3 - 50.3) | 53.0<br>(51.8 - 54.1) |
| **10. Information sent by local (prefecture) government** |  |  |  |  |  |
| Get information: frequently or sometimes | 58.0<br>(57.1 - 58.9) | 53.7<br>(52.4 - 55.0) | 62.4<br>(61.1 - 63.6) | 54.3<br>(52.8 - 55.8) | 60.2<br>(59.1 - 61.4) |
| Trust information: very much or yes | 55.6<br>(54.7 - 56.6) | 52.5<br>(51.2 - 53.8) | 58.8<br>(57.6 - 60.1) | 54.0<br>(52.5 - 55.4) | 56.7<br>(55.5 - 57.8) |

Table 3 shows that almost 90 percent receive information from TV news programs and Internet news sites, and that about 50 percent trust such information. Mainstream scientists are annoyed about the fear-mongering that happens on TV talk and variety shows; indeed, these formats are slightly favored, but considered less credible, among the public. Meanwhile, information from the central and local government (received by 60 percent), including the Prime Minister and the Expert Meeting, is relatively trusted by the participants (50 percent). Among official sources, the local government is most trusted. Newspapers (national and local) are read by only about 42 percent of the participants, and about 48 percent answered that they trust information from newspapers.

Looking at the differences in gender and age, females tend to seek more information and trust it more than males, except for the information from newspapers. Participants over 40 years old tend to access and trust the information from TV, newspapers and officials more than those under 40 years old do, while young people often seek and trust news from the Internet and SNS apps.

### Who does not adhere to social distancing?

As we confirmed in Table 2, more than 80 percent of the participants have been implementing social distancing measures and most Japanese citizens seem to be exhibiting some behavioral change to prevent coronavirus infections. However, this also means that about 20 percent may not be conducting sufficient prevention measures.

To detect what kind of individuals are included in the group not conducting prevention measure, Table 4 shows the estimation results of the logit model. Like the other tables, Table 4 shows the results based on the total, male and female, and under-40 and over-40 categories. The number shown in the table is an odds ratio, so the estimates that are significantly higher than 1 indicate a higher tendency to not conduct proper social distancing.

**Table 4.** **Estimation results of logit model for not conducting social distancing**

| Dependent variable: Avoid places where items 1-3 above overlap in Table 2 -> Not at all or not tue |  |  |  |  | odds ratio, (C.I.) |
| --- | --- | --- | --- | --- | --- |
|  | All | Male | Female | Under 40 y | Over 40 y |
| **Male** | 1.635***<br>(1.304 - 2.049) |  |  | 1.431**<br>(1.039 - 1.970) | 1.854***<br>(1.335 - 2.575) |
| **Age (ref. = 40-49 y)** |  |  |  |  |  |
| 20-29 years old | 1.671***<br>(1.274 - 2.192) | 1.819***<br>(1.285 - 2.574) | 1.647**<br>(1.053 - 2.574) | 1.830***<br>(1.354 - 2.474) |  |
| 30-39 years old | 0.910<br>(0.683 - 1.213) | 0.916<br>(0.639 - 1.314) | 0.949<br>(0.589 - 1.529) |  |  |
| 50-59 years old | 0.819<br>(0.610 - 1.099) | 0.915<br>(0.632 - 1.325) | 0.689<br>(0.423 - 1.123) |  | 0.858<br>(0.634 - 1.163) |
| 60-64 years old | 0.437***<br>(0.264 - 0.724) | 0.631<br>(0.357 - 1.117) | 0.112***<br>(0.026 - 0.471) |  | 0.448***<br>(0.265 - 0.758) |
| **Not married** | 1.445***<br>(1.117 - 1.870) | 1.372*<br>(0.985 - 1.912) | 1.514*<br>(0.979 - 2.340) | 1.476*<br>(0.954 - 2.282) | 1.422**<br>(1.020 - 1.984) |
| **Not having children younger than junior high school age** | 1.246<br>(0.935 - 1.661) | 1.114<br>(0.774 - 1.603) | 1.485<br>(0.922 - 2.392) | 1.366<br>(0.868 - 2.151) | 1.165<br>(0.792 - 1.713) |
| **High or junior high school graduate** | 1.171<br>(0.957 - 1.434) | 1.174<br>(0.914 - 1.508) | 1.152<br>(0.813 - 1.633) | 1.347*<br>(0.999 - 1.815) | 1.053<br>(0.798 - 1.390) |
| **Work status (Ref. = Regular employee)** |  |  |  |  |  |
| Nonregular employee | 1.035<br>(0.795 - 1.349) | 0.992<br>(0.670 - 1.468) | 1.119<br>(0.763 - 1.642) | 0.952<br>(0.654 - 1.387) | 1.151<br>(0.783 - 1.692) |
| Self-employee and others | 0.727<br>(0.446 - 1.185) | 0.958<br>(0.569 - 1.613) | 0.164*<br>(0.022 - 1.197) | 0.658<br>(0.233 - 1.860) | 0.745<br>(0.425 - 1.307) |
| Not working | 0.767<br>(0.552 - 1.066) | 1.089<br>(0.707 - 1.678) | 0.596*<br>(0.343 - 1.035) | 0.745<br>(0.463 - 1.201) | 0.807<br>(0.509 - 1.279) |
| **Household annual income (Ref. = 4,000-5,999K JPY)** |  |  |  |  |  |
| Less than 2,000K JPY | 1.441*<br>(0.990 - 2.099) | 1.103<br>(0.667 - 1.824) | 1.760*<br>(0.970 - 3.195) | 1.883**<br>(1.115 - 3.180) | 1.110<br>(0.638 - 1.931) |
| 2,000-3,999K JPY | 1.159<br>(0.879 - 1.530) | 1.064<br>(0.752 - 1.505) | 1.284<br>(0.797 - 2.068) | 1.480*<br>(0.991 - 2.211) | 0.936<br>(0.632 - 1.385) |
| 6,000-6,999K JPY | 1.182<br>(0.880 - 1.589) | 1.143<br>(0.803 - 1.625) | 1.234<br>(0.713 - 2.137) | 1.430<br>(0.908 - 2.254) | 1.002<br>(0.678 - 1.481) |
| 8,000-8,999K JPY | 0.984<br>(0.687 - 1.411) | 0.893<br>(0.577 - 1.383) | 1.189<br>(0.629 - 2.249) | 1.119<br>(0.633 - 1.977) | 0.877<br>(0.550 - 1.401) |
| 10,000-11,999K JPY | 0.894<br>(0.544 - 1.467) | 0.919<br>(0.523 - 1.614) | 0.737<br>(0.253 - 2.144) | 1.212<br>(0.571 - 2.575) | 0.694<br>(0.357 - 1.350) |
| 12,000-14,999K JPY | 0.730<br>(0.362 - 1.471) | 0.519<br>(0.205 - 1.313) | 1.366<br>(0.463 - 4.029) | 1.748<br>(0.653 - 4.677) | 0.384*<br>(0.137 - 1.079) |
| 15,000-19,999K JPY | 0.460<br>(0.143 - 1.482) | 0.365<br>(0.087 - 1.525) | 0.740<br>(0.097 - 5.656) | 0.415<br>(0.055 - 3.124) | 0.456<br>(0.108 - 1.922) |
| More than 20,000K JPY | 0.707<br>(0.216 - 2.314) | 0.530<br>(0.125 - 2.241) | 2.026<br>(0.246 - 16.664) | 0.434<br>(0.056 - 3.340) | 0.967<br>(0.227 - 4.125) |
| **Drinking habit: Drink 3-6 times per week or everyday** | 1.150<br>(0.911 - 1.451) | 1.278*<br>(0.976 - 1.675) | 0.773<br>(0.476 - 1.256) | 1.326<br>(0.909 - 1.935) | 1.053<br>(0.784 - 1.413) |
| **Smoking habit: Smoke sometimes or everyday** | 1.077<br>(0.856 - 1.355) | 0.956<br>(0.729 - 1.253) | 1.529*<br>(0.995 - 2.350) | 1.124<br>(0.791 - 1.597) | 1.043<br>(0.767 - 1.419) |
| **Big 5 Personality traits** |  |  |  |  |  |
| Extraversion | 1.113***<br>(1.032 - 1.200) | 1.108**<br>(1.008 - 1.219) | 1.125*<br>(0.992 - 1.275) | 1.151**<br>(1.032 - 1.284) | 1.087<br>(0.978 - 1.208) |
| Neuroticism | 0.986<br>(0.909 - 1.069) | 0.977<br>(0.880 - 1.085) | 1.005<br>(0.884 - 1.143) | 0.988<br>(0.880 - 1.109) | 0.987<br>(0.880 - 1.107) |
| Openness | 0.976<br>(0.900 - 1.059) | 0.952<br>(0.861 - 1.054) | 1.014<br>(0.884 - 1.164) | 0.917<br>(0.815 - 1.031) | 1.030<br>(0.919 - 1.154) |
| Conscientiousness | 0.865***<br>(0.798 - 0.937) | 0.865***<br>(0.782 - 0.957) | 0.875*<br>(0.765 - 1.002) | 0.909<br>(0.810 - 1.020) | 0.826***<br>(0.738 - 0.925) |
| Agreeableness | 0.884***<br>(0.814 - 0.959) | 0.906*<br>(0.818 - 1.004) | 0.851**<br>(0.742 - 0.976) | 0.896*<br>(0.799 - 1.005) | 0.874**<br>(0.776 - 0.985) |
| **Constant** | 0.075***<br>(0.037 - 0.152) | 0.138***<br>(0.059 - 0.322) | 0.055***<br>(0.017 - 0.182) | 0.045***<br>(0.016 - 0.123) | 0.100***<br>(0.038 - 0.266) |
| **Number of observations** | 8,548 | 4,679 | 3,869 | 3,021 | 5,527 |

Note: \*\*\* p<0.01, \*\* p<0.05, \* p<0.1

**Table 5.** **“Do you support the government’s policy?”**

|  | % of agree and relatively agree, (C.I.) |  |  |  |  |
| --- | --- | --- | --- | --- | --- |
|  | All | Male | Female | Under 40 y | Over 40 y |
| 1. The government should allow mass gatherings now | 28.8<br>(27.9 - 29.6) | 31.2<br>(30.0 - 32.4) | 26.3<br>(25.1 - 27.5) | 30.5<br>(29.1 - 31.8) | 27.7<br>(26.7 - 28.8) |
| 2. The government should limit movement in addition to mass gatherings | 64.9<br>(64.0 - 65.7) | 64.1<br>(62.9 - 65.4) | 65.6<br>(64.4 - 66.9) | 63.4<br>(62.0 - 64.8) | 65.8<br>(64.7 - 66.9) |

Looking at the estimation results in Table 4, males, people in their 20s and unmarried people exhibit significantly higher odds ratios, indicating that these groups tend not to conduct preventive social distancing. Although work status is not generally associated with this prevention measure, females, regular employees and non-regular employees tended to exhibit higher odds ratios than self-employed or unemployed people.

Regarding household annual income, the lowest group (less than 2,000K JPY) has significantly higher odds ratio for the total, female and under-40 categories.

Higher odds ratios for not conducting social distancing are associated with drinking for males and smoking for females. Furthermore, those with higher extraversion scores also tend to exhibit significantly higher odds ratio in many cases, while conscientiousness and agreeableness are associated with lower odds ratio in most cases.

### Should the government change its policy on mass gatherings?

Before this survey was conducted, the request by the Japanese government for self-restraint in avoiding mass gatherings had become an issue. For example, on 22 March 2020, the K-1 Grand Prix, a martial arts event, was held despite the Minister’s and local governor’s pleas for restraint, and 6,500 participants were packed into the Saitama Super Arena. On 23 March, more than 50,000 gathered in Sendai to see the Olympic flame, which had recently arrived from Greece.^[14]^ We asked the participants whether they supported this policy approach. About 29 percent of participants supported the idea that the government should now allow mass gatherings. Males tend to support allowing mass gatherings more than females. On the other hand, 65 percent supported government limitations of movement, in addition to self-restraint to avoid mass gatherings in order to shorten the period of the pandemic. There are no significant differences in gender and age category for this question.

## Discussion

Under circumstances where there is no enforced ban on mass gathering or travelling beyond the home region, our findings indicate that a large portion of Japanese citizens seem to be implementing proper prevention measures on their own.

First, more than three-quarters of the survey participants have taken some preventive actions, including social distancing, handwashing, coughing etiquette and strengthening immunity. Because the previous empirical studies did not include developed countries like Japan, ^[15]^ there is little scientific evidence that Japanese people prefer cleanliness and tend to wash their hands relatively more frequently than other countries. In Japanese communities, water facilities for handwashing with soap and hand sanitizers are normally placed various public places, such as train stations and supermarkets. Moreover, handwashing became part of regular practice at home and school through post-war education.^[16]^ In general, Japanese people have developed the discipline of washing their hands before having meals or after using the toilet. It is also well known that Japanese bow for greetings instead of handshaking, kissing and hugging. This cultural behavior implies that the frequency of body contact among Japanese people may be less than for those who greet with handshaking, kissing and hugging. During hay fever season, Japanese citizens regularly wear surgical-style masks for prevention; it may be that wearing a mask was a less popular preventive measure than some of the others in this study due to there being a shortage of these products. These already habitual practices might help behavioral changes develop among Japanese citizens during these unusual times.

In the survey, more than half of the participants had not prepared access to consultation centers or transportation methods for if they were to become ill, implying that they had not planned for the possibility of contracting COVID-19. We must advise the public to prepare for such an event, to talk to family and close friends about unexpected advanced care planning, and to imagine not being able to use ventilators or extracorporeal membrane oxygenation at the severe stage.

Second, one of the main motivations for people’s behavioral changes was the infection aboard the *Diamond Princess* cruise ship in early February 2020. At that time, only a few cases of domestic infection had been reported in Japan, but news of the quarantine and positive test results among the passengers was broadcasted daily. This may have contributed to Japanese citizens changing their mindset and behavior toward precautionary measures earlier than in Europe and the US. The sudden request by the Prime Minister for nationwide school closures in the end of February might also have been an effective measure for changing the mindsets of Japanese citizens toward prevention, even though this move was scientifically questioned and confusing to the public, especially to single parents and double-income households.

Third, information from the Expert Meeting and central/local governments, including the Prime Minister, are relatively trusted by survey participants. The Expert Meeting and central/local government have held frequent press conferences, trying to clarify the tentative scientific risks and advocating for citizens to conduct prevention measures. Such crisis communication attempts may have caused behavioral changes in Japanese citizens. The most trusted resource in this study was information from the local government, which was a hopeful result, considering that the countermeasures against the virus are decided and conducted by the local government. It is also important to support residents with disadvantages to information and communication in the local communities.

However, the results also show that about 20 percent of the participants are reluctant to implement proper prevention measures. The statistical analysis indicates that typically those people are male, younger (under 30 years old), unmarried, are in lower income household, have a drinking or smoking habit and have a higher extraversion score. To prevent the spread of infection in Japan, it is imperative to address these individuals and encourage their behavioral change in various ways that will reach and move them. It is notable that approximately 65 percent of the participants support stricter countermeasures, such as limitation of movement. As we mentioned in the introduction, as of yet, the government has not made stay-at-home orders mandatory and it has not offered financial aid to those affected by such measures. The current requests from central/local governments are not legally binding and people/businesses have to arrange financial compensation independent of the government. We should observe how effective these measures are in Japan over the long term.

There are several limitations to this study. First, the data were self-reported, and participants’ actual behaviors have not been observed. Second, the sample was not collected based on random sampling. Quota sampling ensured a similar distribution to the Japanese population among demographic groups (gender, age and work status), but the sample within each group does not necessarily reflect the population. Third, we obtained this dataset at the end of March 2020, when the infection is not explosively widespread in Japan. This study should be reanalyzed after the COVID-19 pandemic comes to an end.

## Data Availability

All relevant data are deposited in openICPSR.

https://doi.org/10.3886/E118584V1

## Acknowledgments

We would like to thank the participants in our online survey for offering valuable data. This work was supported by university grants allocated to the Department of Public Policy, Human Genome Center, The Institute of Medical Sciences, The University of Tokyo. We also appreciate members of the COVID-PAGE (Public Advisory Group of Experts) for insightful discussions.

## Notes

### Competing Interest Statement

The authors have declared no competing interest.

